# Computer Vision Estimation of Stress and Anxiety Using a Gamified Mobile-based Ecological Momentary Assessment and Deep Learning: Research Protocol

**DOI:** 10.1101/2023.04.28.23289168

**Authors:** Ali Kargarandehkordi, Peter Washington

## Abstract

Stress and anxiety can contribute to the development of major health problems such as heart disease, depression, and obesity. Due to its subjective nature, it is challenging to precisely measure human affect by relying on automated approaches. We therefore propose a personalized prediction framework fine-tuned for each participant in lieu of the traditional “one-size-fits-all” machine learning approach. We aim to collect such individualized data via two distinct procedures: 1) a smartphone-based ecological momentary assessment of stress, and 2) Zoom calls. The data collected from these periodic self-reports will include selfie photographs and ecological momentary assessments of affect. To enhance user engagement during the data collection process, we propose the use of gamification, an emerging trend which involves influencing user behavior and lifestyle by incorporating fun and engaging game elements into non-game contexts (e.g., health-related tasks). In addition to developing a standardized platform to achieve more accurate recognition of stress and anxiety, we plan to conduct a concurrent study in which we will capture videos of our subjects undertaking the Stroop Color Word and Amygdala Test and analyze the footage to identify additional significant characteristics that relate to anxiety. This could include features such as head and mouth movements, lip and cheek deformations, eye gaze, and blinking rates. The final results will provide a comparative evaluation of both objective measures of stress. This research project was approved by the University of Hawaii Institutional Review Board.

## Introduction

Consistent exposure to stress and repetitive feelings of anxiety can lead to detrimental impacts on life quality and expectancy [1]. Several of the leading causes of death (e.g., cardiovascular disease) are linked to these adverse mental states [2, 3]. Despite this large burden on the health system, chronic stress and anxiety are largely left undertreated [4, 5]. The recognition of stress and anxiety is a highly subjective endeavor, leading to challenges with quantifying such affects in an objective manner. Generally, stress and anxiety are measured through clinical questionnaires designed by psychological experts [6].

In certain cases, stress and anxiety have been documented to produce a noticeable effect on facial characteristics, including modifications on pupil size, eyelid closure, blinking rate, mouth activity, lip deformation, cheek deformation, and head movement[7-9]. However, this physical manifestation varies significantly across individuals. Therefore, developing an accurate single-purpose machine learning (ML) model to recognize stress and anxiety is likely outside of the realm of feasibility.

To overcome this challenge, we propose the utilization of a personalized ML approach, where an individual ML model is fine-tuned to each individual participant’s data [10, 11]. Such personalization can provide an opportunity to eventually develop digital therapeutics which intervene when stress or anxiety is recognized.

In this protocol documentation, we first present a survey of methods in the field of digital mental health. We then discuss a data collection protocol for curating a dataset of facial selfies and corresponding labels of stress and anxiety through an ecological momentary assessment (EMA). We next discuss a series of personalized ML experiments which we will conduct using the resulting dataset.

## Prior Works

### Conventional Methods for Stress and Anxiety Measurement

There are two commonly used methods to assess anxiety disorders: (1) analyzing data collected through clinician interviews and (2) self-reports using standardized questionnaires [12]. These methods involve asking patients to report on the severity and intensity of their anxiety symptoms. However, these measurements are subjective and may result in inconsistent or unreliable data [13] due to numerous factors including the patient’s inability to recall information accurately and discomfort with the clinician [14]. Other factors that can compromise data reliability include question ambiguity, psychopathology, and biases [15, 16].

Another way to measure stress and anxiety is to evaluate cortisol levels produced by the HPA axis. This involves an intensive analyzation of cortisol from different sources such as blood, urine, hair, and saliva [17, 18]. Other physiological indicators include assessing brain activity using EEG, heart activity using ECG, muscle activity using EMG, and skin conductance using electrodermal response. Changes in breathing patterns and speech characteristics can also indicate anxiety [6, 19, 20]. Researchers are particularly interested in changes in fundamental frequencies of the voice as a potential indicator of anxiety [7, 21].

### Facial Signs and Expressions

Charles Darwin’s book “The Expression of the Emotions in Man and Animals” in the late 1800s first highlighted the connection between emotional states and facial expressions [22]. Later studies in the 1900s and early 2000s provided more evidence to support this connection, particularly with advanced methods like HPA axis and cardiovascular responses to stress. Recent studies suggest that facial signs can provide insights into classifying anxiety[7, 8]. Anxiety can affect various facial expressions including eyes (pupil size variation, eye aperture, blinking rate, and gaze distribution), mouth activity [23], specific deformations on the lips [24], cheeks, head movements and velocity [25, 26]. However, covarying factors such as lying, depression, Parkinson’s [27], schizophrenia [28], and environmental conditions like lighting and humidity can also affect facial expressions, particularly blinking rates [29].

### Machine Learning for Affective Computing

Machine learning (ML) models are increasingly applied to healthcare for various purposes such as therapy planning, disease diagnosis, and illness prediction. These tools have been shown to improve the quality and efficiency of medical care and are considered promising for medical experts [30, 31]. Although machine learning models have been successfully used in mental health to predict psychological and perceived anxiety, there are challenges related to reliability due to limitations in data collection [32], limited research-only resources [33], confidentiality of the data and inability to disclose it [34], and lack of effective commercial licenses [35]. These challenges make it difficult to use machine learning models in real healthcare scenarios.

Due to the inherent heterogeneity and subjectivity of stress, recognizing stress and anxiety typically involves using multiple forms of data. Analyzing and merging these multimodal data can help create a reliable and stable stress and anxiety recognition models. There are various ways to measure stress, including using stressors like personal factors, environment, and individuals’ physiological, psychological, and physical reactions to stressors. Eye data and vehicle dynamics data are easily accessible, and some researchers have explored their connection to stress [36]. Past research has mostly used conventional machine learning techniques to manually extract features from data and create stress recognition models. Although this method has yielded favorable results, it can be difficult to accurately identify important and representative features, and it requires specialized knowledge and is less resilient to noise and data variations [37]. Some researchers have developed stress level recognition models using multimodal data. Benoit et al. [38] developed a driver simulator that utilized video data (facial activity) and physiological data, such as blinking, yawning, and head turning, as well as ECG and electrical skin responses to measure the driver’s stress levels. Rigas et al. [39] developed a model that used physiological signals, video recordings (eye data, head movement), and environmental data to classify driver stress levels and predict driving performance. The model achieved an 86% accuracy rate in distinguishing between two stress levels (no stress, stress) using an SVM classifier.

Generalized machine learning models have limitations in recognizing emotional states due to high variability in physical and personal characteristics among individuals, leading to poor performance [40]. Personalized machine learning, by contrast, has been suggested as a more effective technique for precision health machine learning with high-dimensional data on subjective and person-specific prediction classes [40, 41]. This approach involves further research on affective variations within the individual, where each model is trained and tested solely on an individual’s data to examine the effects of variability on recognizing emotional transitions and states.

### Smart-sensing and Passive Monitoring

Observing and evaluating patients’ behavior as an indicator of changes in the severity levels of mental disorder symptoms can be an alternative to traditional methods. Using mobile phones as sensing devices enables researchers to passively record specific behaviors related to different mental disorders. Mobile phones are useful tools due to their various sensors and software for evaluating behaviors and contextual awareness as well as their constant presence with the owner [42-44]. In addition to subjective information from interviews or self-reports, mobile phones can also provide objective data through their camera, allowing researchers to make objective assessments of mental health [45].

### Reminders and Alerts

Adding alert-based components such as reminders to digital health interventions can improve engagement and encourage adherence to the intervention. These reminders can be sent through various channels, including email, SMS, or push notifications within mobile applications. Incorporating reminders has been shown to increase engagement levels in mental healthcare research, making it more effective than interventions without this feature [46-48]. Using reminders in digital health interventions can also lead to higher levels of adherence to the research and lower non-usage attrition, which are common issues with online-based interventions [49-51]. Non-usage attrition refers to participants who stop using the digital intervention while still participating in the research through other protocols, such as filling out questionnaires.

### Gamification

Gamification, which involves using game design elements in non-game contexts, is another popular trend to improve the user engagement and activities in digital interventions. In an academic context, gamification refers to using gamified elements and incentives, along with visually appealing designs, to enhance user engagement, motivation, and adherence to research activities [52-54]. Gamification has been implemented in various healthcare services to increase user activity and productivity of behaviors, using different elements such as points, leaderboards, challenges, feedback [55-60], rewards, story/theme [61, 62], achievements/badges [55], progress tracking, and levels [58, 60].

### Study Procedure and Methodologies

The Ecological Momentary Assessment (EMA), also known as the Experience Sample Method or Ambulatory Assessment, is a useful tool for classifying momentary moods in natural and real-life environments [63-65]. This method allows individuals to report their current mental state in real and familiar environments rather than in unfamiliar settings through standardized instruments [65-67]. EMA involves regular self-reporting of a psychiatric property while carrying out daily tasks over a specified period of time. Digital healthcare interventions can utilize EMA, which has methodological strengths including increased ecological validity, reduced retrospective bias and errors, and the ability to collect data periodically across different real-life situations [65].

This study will use both a smartphone-based EMA application (which we call STAND) and Zoom calls to measure stress. The first procedure involves assessing stress levels by asking study participants to take daily selfies and self-report their stress and anxiety levels as well as a description of how they feel at the moment (including any symptoms related to stress/anxiety). In this procedure, the data collected will consist of both images and text, which we will use to train single modality and multimodal deep learning models for predicting stress and anxiety levels. We will evaluate the extent to which each modality improves the accuracy of the prediction. The deep learning CNN/LSTM network architecture we intend to employ for training the model in the first procedure is depicted in Figure 2.

**Figure 1:**
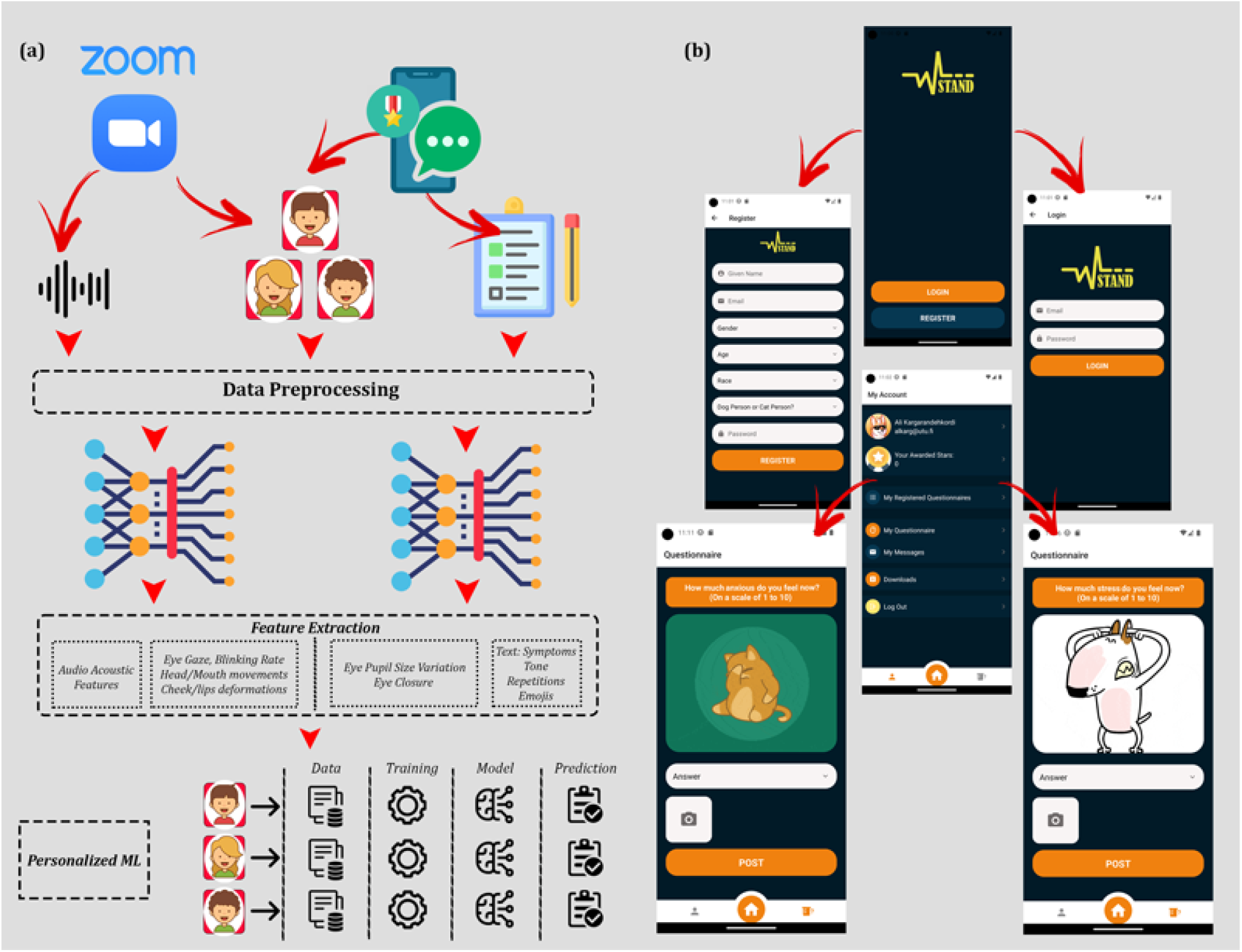
a) Workflow of the study. b) Flow of different screens in the STAND app.

**Figure 2:**
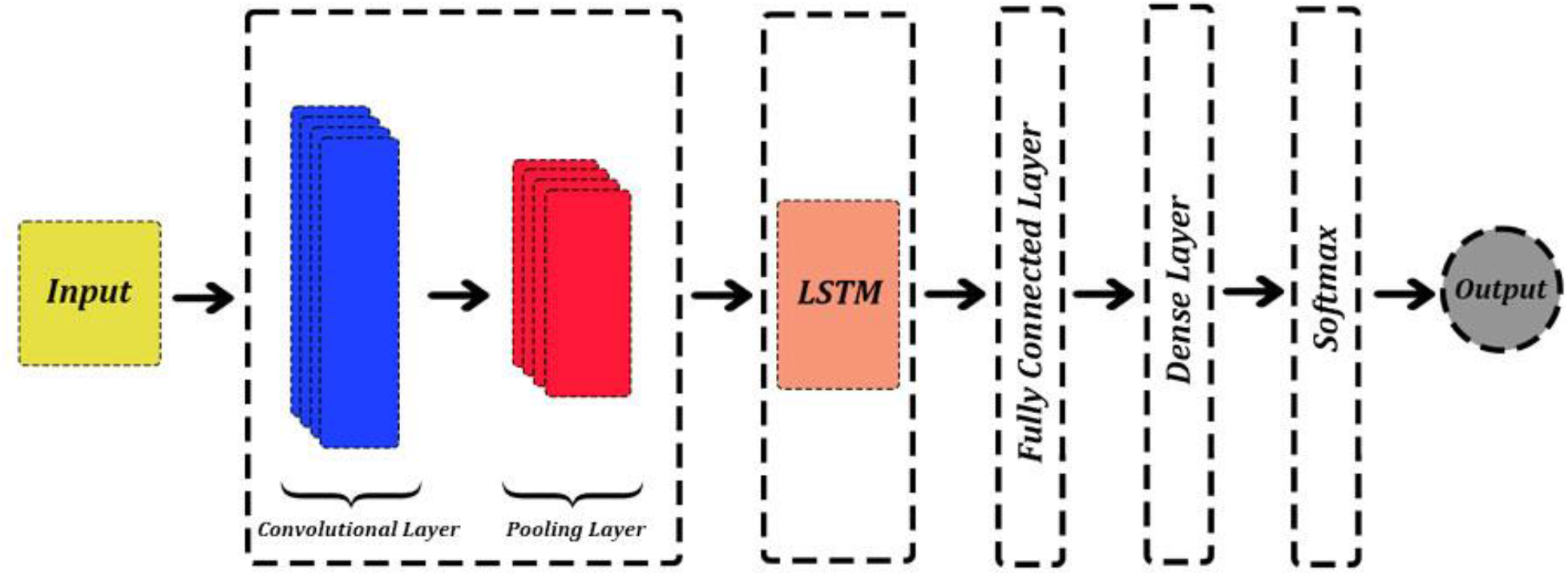
Neural network architecture of the proposed single modality CNN/LSTM network.

During the second phase of the study, participants will be required to participate in periodic Zoom meetings at predetermined intervals throughout the one-month data collection period. These meetings will involve engaging in psychological tests such as the “Stroop Color Word Test” and “Amygdala Test,” which are designed to be game-like in nature. Participants will be recorded during these tests, and the videos will be carefully analyzed to identify any additional facial expressions that may be associated with stress and anxiety.

Based on an EMA based intervention, this study intends to utilize all the techniques mentioned in the Prior Works section to efficiently recognize stress, a commonly experienced emotional state. The study will involve collecting data from participants for a month and then analyzing the data to identify the key features associated with accurately recognizing stress and anxiety. The experiment will be conducted in three phases, which are shown in Figure 1. Advantages of using an EMA-based mobile application for digital intervention in the healthcare field include greater availability, accessibility, anonymity, flexibility, cost-effectiveness, and interactivity through visually attractive designs [68-70]. We aim to explore the impact of incorporating personalized app modifications such as animations and informal language in increasing user engagement with the digital intervention. An example of this is using an interactive question related to a user’s preference for dogs or cats as part of the app’s registrations process (a dog person, or a cat person?). The app selects and arranges animations in various sections based on the user’s response to this question.

Under an expedited review procedure, this research project was approved on December 20, 2022, by the University of Hawaii Institutional Review Board (UH IRB). The application qualified for expedited review under 45 CFR 46.110, Category 6, 7. Per 45 CFR 46.109. This study targets students as the main subject for analysis and focuses to screen stress among this population. Data associated with the students are prone to challenges resulting from barriers such as lack of time, stigma, scheduling issues, and significant workloads [71]. Stress is among the major pervasive mental health concerns within the school community, and many students around the globe undergo increased levels of stress in their everyday life [72, 73]. We aim to recruit at least 20 students (gender balanced) aged 28 ± 10 years.

### Stage 1: Mobile Application

A mobile application that can work on both Android and iOS platforms has been developed for the purpose of investigating the development of a subjective measure of stress. Study participants will receive questionnaires through daily notifications on the STAND app, in which there is an option for taking selfies. The photographs will be used to analyze potential stress related facial expressions (objective measurement). Initially, users will receive two separate notifications throughout the day that inquire about their levels of stress and anxiety. There will be a space provided where participants can input their current emotions and any related symptoms they may be experiencing such as tiredness, butterfly in the stomach, lack of focus etc.

To introduce game-like aspects to the study, a system of rewarding stars has been included in the app. The participants will be awarded stars according to the number of points they obtained for their enthusiastic and constructive involvement during the data gathering stage, such as completing assignments within 30 minutes of receiving a notification and providing complete responses to all questions each day during the initial phase. Participants who earn more stars will have a higher chance of winning rewards, such as grocery shopping gift cards, through a fair lottery drawing at the end of the data collection phase.

### Stage 2: Ecological Momentary Assessment Study and Periodic Zoom Calls

In the second stage of the study, the process mentioned above will be carried out for one month to gather adequate data needed for creating personalized machine learning algorithms. To evaluate relevant psychometric properties in the third stage, participants will be requested to complete a self-report questionnaire every day. The questionnaire consists of two questions and will be distributed as multiple notifications throughout the day, with each notification containing one question. Every time participants receive a notification; they will need to respond to the same questions they have answered in the previous days on a scale ranging from 1 to 10:

- How much stress do you feel right now?
- How much anxiety do you feel right now?

In a second concurrent procedure, participants will also attend periodic Zoom meetings (at fixed intervals within the one-month data collection experiment) to take part in a game-like psychological examination known as “Stroop Color Word Test” and “Amygdala Test” while they are recorded. The recorded videos will be scrutinized to identify additional facial expressions associated with stress such as eye gaze distribution, blinking rates, lips and cheeks deformations, mouth activity, and head movements. Extracted audio signals from recorded videos will also be analyzed alongside facial expressions in a multi modal deep learning network (Figure 3).

**Figure 3:**
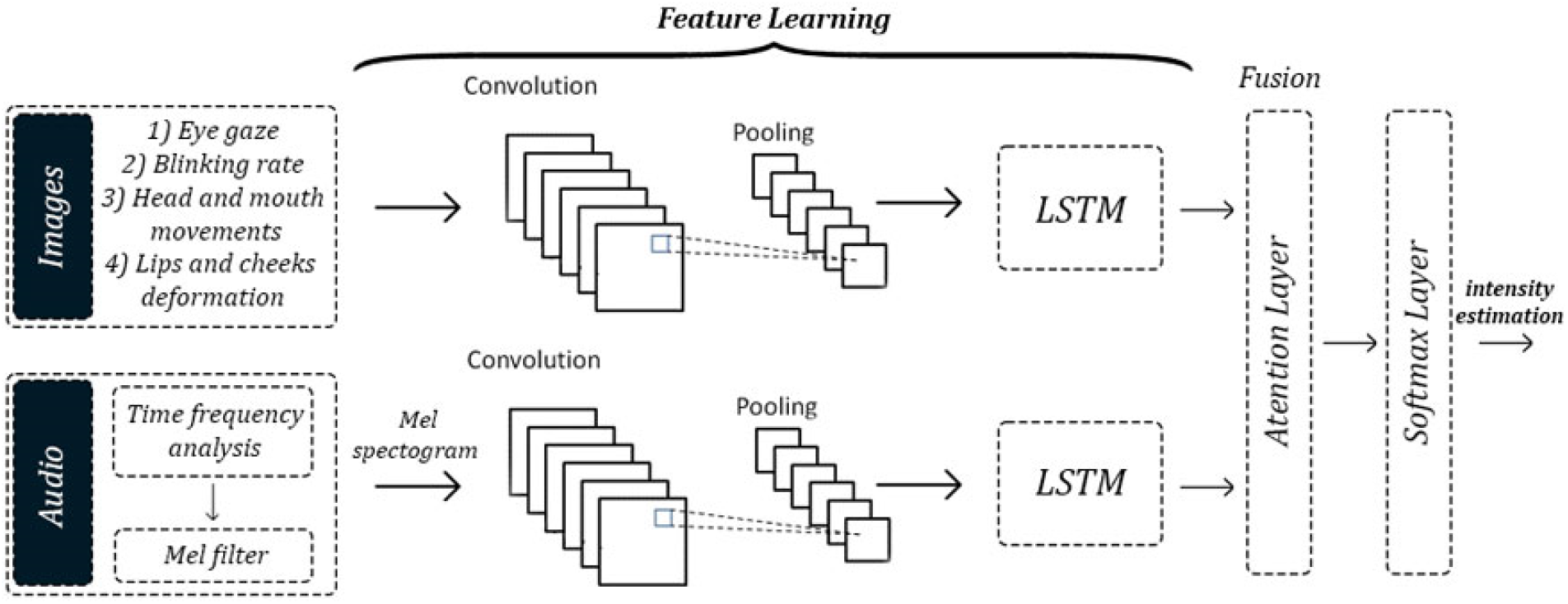
Neural network architecture of the proposed CNN/LSTM multimodal network.

During this process, individuals who have already been diagnosed with anxiety will be requested to take part in the two psychological tests mentioned earlier. The amygdala test requires the participants to search through arrays of faces with a specific emotional intensity level. Previous research has suggested that anxious individuals are quicker to detect angry facial expressions than positive ones. [74]. Therefore, individuals with anxiety tend to exhibit an increased attentional bias towards cues that indicate any form of threat in comparison to individuals without anxiety. [75]

### Stage 3: Deep Phenotyping of Stress and Anxiety

In the third and final phase, we will train personalized deep learning methods to recognize stress and anxiety. During the first procedure, two modalities will be utilized both individually and concurrently via separate network architectures to determine the marginal impact of each modality on prediction accuracy. These modalities consist of facial expressions taken from daily selfies and text descriptions provided by the participants concerning their momentary feeling. The overall CNN/LSTM network architecture of the model that will be employed to train each modality is displayed in Figure 2. Alongside a convolutional network, LSTMs [76] will be used to capture temporal relationships. LSTMs are useful for capturing long-range dependencies in input sequences such as videos [77]. The LSTM’s output will be transmitted into a fully-connected network layer, which will then be transmitted into another dense layer. This dense layer will then be forwarded to a final dense layer that utilizes softmax activation to estimate stress and anxiety levels.

In addition to an enhanced subjective and objective measurement of stress (first procedure), this work presents a method for recognizing stress in students using a personalized deep learning framework that incorporates audio and facial expressions extracted from Zoom calls to predict self-reported stress and anxiety. The proposed multimodal framework involves four main steps: preprocessing, feature extraction, feature fusion, and classification (Figure 3).

The model will extract stress-related information by fusing data from multiple sources, including facial expressions (e.g., eye, head, cheek, mouth, and lips features), and audio signals. The voice data will be transformed into a time-frequency representation, and the facial expressions are extracted from photographs and video frames. The multidimensional features of each modality will then be extracted using deep learning techniques. The fully connected layers in the framework obtain information about the stress state, which is fused into a global matrix for stress and anxiety recognition and intensity level. The ResNet50 and I3D models will be used for audio and facial expression feature extraction, respectively, with the I3D model incorporating a temporal attention module to identify important frames. The proposed multimodal fusion model aims to classify stress and anxiety levels in students in real time using multiple sources of data.

This study features a personalized analysis approach that utilizes deep learning to learn different features associated with a person’s subjective symptoms, head and mouth movements, deformations of cheeks and lips, eye gaze, variations in pupil size, and blink rate. This approach enables the models to be overfitted to individual human subjects (i.e., “personalized machine learning”) by self-supervised learning. By analyzing the top predictors in each individual, we can identify the distinct features that contribute to their stress and anxiety over time. All the features used in the personalized models can be targeted for intervention. Therefore, the results of the personalized models can be used to create individualized interventions with a single or multiple feature-based approach, which can be used to recognize stress and anxiety levels using a personalized multimodal strategy. This method creates a unique model for each patient that is specifically designed to classify their stress and anxiety levels, and it will only be used for that particular patient when deployed into clinical contexts.

## Conclusion

Previous research studies have shown that mental health applications can be an effective tool for increasing user engagement even without incorporating gamification into their design. We anticipate that the STAND app will result in greater user participation. It will be beneficial to explore the addition of more personalized and visually appealing design elements. By incorporating a second concurrent procedure into this research protocol and analyzing video frames facial expression and audio signals, we will have the opportunity to examine and compare the performance of our smartphone-based EMA application with another robust procedure that both can objectively measure the intensity of stress and anxiety. The use of personalized machine learning techniques in digital health has unlocked promising opportunities for creating and optimizing models in the field of mental health diagnosis, management, and treatment. The results of this study will compare the accuracy of both subjective and objective measurements of stress and anxiety intensity, which can serve as a resource for future studies.

## Discussion and Future Works

The proposed study aims to provide evidence for the efficacy of using state-of-the-art technologies (i.e., ecological momentary assessment and personalized machine learning) to classify stress and anxiety levels among young adults. Combining these technologies to make digital mental health interventions might be a promising alternative to the current standard of care. Although we will apply novel techniques to quantify anxiety and identify the potential relationships between facial cues for anxiety and stress, there are still a few technologies that need to be investigated and combined in future works to enhance the efficiency of such recognition mechanisms.

Crowdsourcing is one of these technologies that has emerged in the field of digital phenotyping. Judging and validating subjective details often requires human intervention for potential improvements. Innovations in this field include algorithms for data efficiency and complexity management [78, 79] and debugging and verification of AI models [80, 81].

Another solution to increase the efficiency of mental health interventions lies in modern smartphones and their sensors. The prevalent usage of smartphones in society, and the fact that they became an integral part of individuals’ lives represents a new frontier technology in digital health. Physical activities approximated by acceleration sensors[82], duration of stays and their specific time of the day at known locations approximated by Wi-Fi and GPS information [83], and the frequencies of interactions and social activities [84] can be highly related to the individual’s mental health state and can be screened and recorded passively benefiting from these portable devices.

Our plan is to develop the STAND app into a versatile tool that can be customized for researching and gathering computer vision data of the face to build personalized machine learning models which can support digital health interventions related to mental states and developmental disorders. This work can integrate with existing research in using automatic emotion recognition for a variety of contexts, including mental illness diagnosis, recognizing human social and physiological interactions, and developing sociable robotics and other human-computer interaction systems [57, 85-89]. For example, emotional expressions have a crucial role in recognizing certain types of developmental disorders. Autism spectrum disorder (ASD) affects almost 1 in 44 people in America [90] and it is the fastest-growing developmental disorder in the United States [91, 92]. Children with autism exhibit emotions differently than their neurotypical peers, and they often struggle to display appropriate facial expressions. [93-95]. In order to enhance social communication, it is anticipated that creating an optimal model for human-computer interactions would involve accurately classifying and responding to human emotions e.g., stress and anxiety [96-105]. Initial attempts have been made to utilize digital and wearable devices to enable families to conduct therapy sessions in the comfort of their own home. The intervention structure can be tailored to meet the specific needs of the child [106-112]. The personalized affective AI proposed here could be integrated into these interventions to provide enhanced digital and mobile therapies to children with ASD and other developmental delays.

## Data Availability

The dataset will be stored on cloud storage provided by Amazon Web Services (AWS) with extra security measures and might be published online for future use in the analysis of potential facial expressions in other AI studies and research. This includes the right to edit or duplicate any images/recordings;

